# Will online NHS 111 reduce demand for the telephone NHS 111 service? Mixed methods study of user and staff views

**DOI:** 10.1101/2022.11.29.22282892

**Authors:** FC Sampson, EL Knowles, J Long, J Turner, JE Coster

## Abstract

**Introduction:** Online NHS111 was introduced in 2018 in response to increasing and unsustainable demand for Telephone NHS111. We explored user and staff perspectives of telephone and online NHS111 to understand how the two services were used, and whether and how online NHS111 had potential to reduce demand for telephone NHS111.

**Methods:** We used a convergent parallel mixed methods design, using data from the national online NHS 111 user survey and telephone user survey for 2 NHS 111 areas and semi-structured interviews with 32 recent users of online 111 and 16 NHS 111 staff. We analysed survey data for 3728 online users and 795 telephone users in SPSS, using chi-squared test for proportions and adjusting for age, sex, ethnicity and presence of long-term conditions. Qualitative data was analysed using Framework Analysis.

**Results:** Telephone NHS111 health adviser skills in probing and obtaining ‘soft information’ were key to obtaining advice that was considered more appropriate and trusted than advice from online interactions, which relied on over-simplified or inappropriate questions. Telephone users were more satisfied with NHS111 than online users for all comparable measures, reported higher compliance with advice and were more likely to say they would have contacted another service if they hadn’t used NHS111 (p<0.001).

Online NHS111 was perceived to provide a useful and convenient adjunct to the telephone service and widened access to NHS111 services for some subgroups of users who would not otherwise access the telephone service (e.g. communication barriers, social anxiety), or were concerned about ‘bothering’ a health professional. The nature of the online consultation meant that online NHS111 was perceived as more disposable and used more speculatively.

**Conclusion:** Online 111 was perceived as a useful adjunct but not replacement for telephone NHS 111 with potential for channel shift hindered by reduced confidence in the online service.

**What is already known on this topic:**

- Online NHS111 was introduced in the UK in 2018 to reduce unsustainable demand on the NHS111 telephone service.
- Quantitative routine data analysis showed that the introduction of online NHS111 had limited impact on demand for the NHS111 telephone service but does not explain how or why ‘channel shift’ of demand from the NHS111 telephone service may not be happening.

*What this study adds:*

- Users trusted and followed advice from telephone NHS111 more than online NHS111 due to the human interactions involved in answering questions appropriately.
- Online NHS111 was used more speculatively and advice potentially seen as more ‘disposable’ due to the lack of contact with health professionals.
- The introduction of online NHS111 improved overall access to NHS111 services for a subset of users. How this study might affect research, policy, practice

- Online triage has limited potential for shifting demand due to ambiguity in algorithm question wording.
- Refinement of questioning will be required for online NHS 111 to increase in value and use for people with multimorbidity and long-term conditions. A hybrid option whereby online users can clarify question meaning using live chat options may improve the usefulness of online NHS111.

## Introduction

In the UK, emergency and urgent care is provided by a range of services including emergency services (999 ambulance service, emergency departments), urgent care services (out-of-hours primary care, minor injury units, walk-in centres, urgent care treatment centres, NHS 111) and in-hours primary care (requests for same day appointments and telephone advice). There is widespread concern about rising and unsustainable demand for urgent and emergency care (UEC) services, which are not wholly explained by population increases or an aging population. (1-3) The NHS 111 telephone service (Tel111) was set up to provide triage and assessment and to direct people with low acuity non-emergency health problems to the most appropriate UEC service, or provide self-care advice. Health advisers (usually non-clinical) ask the caller a number of questions, following different clinical algorithms, and then provide a clinical ‘disposition’ or recommendation about what service to contact, and the time period for doing so. For some callers a transfer to, or callback from, an NHS clinician will be offered.

However, demand for the Tel111 service has risen significantly, with nearly 15 million calls in 2017/18 (4), resulting in the service struggling to meet demand and an increase in waiting times for urgent calls. In an attempt to stem the increasing demand for Tel111, NHS England rolled out online NHS 111 (OL111), a digital alternative to Tel111 in 2018. (1, 5) OL111 uses the same clinical algorithms as Tel111 but requires users to direct themselves through the initial questions using a computer, tablet or smartphone. Users of the online service will receive a recommendation (disposition) advising them what service to use, providing details of local services and availability, or may get a call back from a clinician. At the time of rollout, little was known about how acceptable the online service would be to users, and whether there was potential for OL111 to offset or ‘channel shift’ demand for Tel111 (i.e. patients moving from use of Tel111 to OL111).

There is some evidence that the user profile of digital and online symptom checkers and health assessment services differ from other platforms (online users being more likely to be young, employed and female). (6-8) This suggests that OL111 may reach a different audience but not channel shift, and the launch of OL111 was accompanied by concerns of induced demand rather than a shift in demand. (9) The process and outcomes of the OL111 service may lead to different user experience, with evidence suggesting other algorithm-based triage systems to be more risk-averse than those involving healthcare professionals. (9, 10)

As part of a wider evaluation of the impact of the introduction of online NHS 111 on the NHS 111 telephone service (11), we explored user and staff perspectives to understand awareness and acceptability of the online service, how the service is used and whether there is potential for channel shift from Tel111 to OL111. Within this paper we explore perspectives of how and why people use the two services differently in order to understand the potential for channel shift from a telephone to an online service.

## Methods

We used a mixed-methods approach to understand the acceptability and usability of OL NHS 111, explore potential for ‘channel shift’ and other impact on telephone service. Due to short overall project deadlines, we used a convergent parallel design with quantitative and qualitative elements undertaken concurrently, different components analysed independently then results interpreted together. (12) We incorporated three different data sources: staff and stakeholder interviews, survey of online and telephone NHS 111 users and NHS 111 online user interviews. Equal weight was given to quantitative and qualitative data and we compared the results of the different datasets to understand where results supported or contradicted each other. We present the ‘converged’ results from our side-by-side analysis of the different data sets. We recruited 4 NHS 111 case study sites to provide staff and Tel111 user data, although only two sites were able to provide Tel111 user data during the timescale of the project.

### Staff interviews

Recruitment took place between November 2019 and June 2020. Information about the study was circulated to relevant staff via key contacts at each site, and participants were asked to contact the research team directly if they wished to take part. We aimed to purposively sample staff with a variety of roles and length of experience. After a low initial response rate, we amended the process to offer participants a shopping voucher and asked sites to recirculate the invitation. Recruitment continued to be low due to significant pressures on the service and was then halted due to the Covid-19 pandemic.

Interviews were conducted by telephone by one member of staff (FS) and digitally recorded. Interviews lasted an average of 33 minutes (range 15-62), and were transcribed verbatim. Data were loaded into NVivo software to organise thematic analysis. The coding framework was developed by the interviewer in consultation with two colleagues, who also read a sample of transcripts. The framework was based upon a priori themes including resource implications, perceptions of use of NHS 111 online and role of the service within UEC system. We developed additional themes around trustworthiness and belief, knowledge and awareness, and integration.

### Survey of NHS 111 online and NHS 111 telephone users

We used pre-existing user experience datasets from providers of NHS111 online and telephone services, requesting key data questions and asking providers to amend questions for our purposes where possible in order to obtain similar variables. NHS111 Online user experience data for all users was obtained from NHS Digital between September 2019 and May 2020. NHS111 Telephone user experience data was obtained for two of the case study sites between October 2019 and February 2020, provided by the ambulance services. Most questions used closed response sets, with some allowing for additional free text comments.

Users of NHS 111 Online were invited by NHS Digital to complete a questionnaire directly after their assessment, and to provide their email address if they were willing to receive a second survey approximately two weeks later. Telephone users received a postal questionnaire, sometimes a number of weeks after their contact with the service, which may have implications for recall. All data was collected by the service providers and anonymised before being forwarded to the research team.

We undertook descriptive analysis of NHS 111 Online and NHS 111 Telephone user data using IBM SPSS Statistics version 26. We used binary logistic regression and multinomial logistic regression to adjust for age, sex, ethnicity, and presence of a long-term condition. NHS Digital provided anonymised free text comments from NHS 111 Online respondents in our initial case study areas. We read these free text comments and grouped the data into themes.

### Online user interviews

After completion of second NHS Digital survey participants could view a link inviting them to take part in an interview and provide their contact details. A researcher then contacted individuals by telephone and arranged a time and date for the interview as soon after contact as possible to ensure good recall of events. We planned to undertake up to 40 interviews. We initially took a non-selective approach to sampling, which resulted in a largely female, middle aged, ethnically white sample, who had received a call-back from NHS 111 after their online assessment. After completing 20 interviews, we purposively recruited only younger service users and those who had not received a call-back.

Semi-structured interviews took place by telephone between December 2019 and April 2020. Informed consent was recorded at the start of each interview. The topic guide covered general use of online services and health services, previous use of NHS 111 (telephone and online), experience of using NHS111 Online, any preferences between the online service and telephone service, and the likelihood of using the online service in the future. One researcher (JL) undertook all of the interviews, which lasted between 24 and 55 minutes. Participants were sent a shopping voucher on completion of the interview.

Interviews were audio recorded and transcribed verbatim. Transcripts were analysed in NVivo using framework analysis. The framework was developed from our research questions, the topic guide and an initial reading of a sample of transcripts. Themes included positive and negative aspects of the experience of NHS 111 Online, compliance with advice, propensity to use NHS111 Online again, and recommended future development of the service. Once the initial framework was agreed, the researcher who had undertaken the interviews coded all transcripts to the framework, adding further emergent themes and sub-themes and discussing any areas of uncertainty with a second researcher. These two researchers then read extracts within each theme to refine the final thematic framework

### PPI

We used an established Patient and Public Involvement (PPI) user group to support the study, including one member who was a co-applicant on the research grant and attended project management group meetings throughout. The group advised on documents related to ethical approval, discussions of emerging themes within analysis and discussions of early drafts of results.

## Results

### Characteristics of respondents for online and telephone 111 users

Telephone respondents were more likely to be older, male, white British and less likely to have a long term condition than online respondents. Interview participants were broadly representative of the online user population.

|  | **Telephone survey** | **Online survey** | **p-value** | **Interviews** |
| --- | --- | --- | --- | --- |
| < 35 | 17 (118) | 21 (755) | <0.001 | 22 (7) |
| 35-65 | 32 (227) | 55 (2001) |  | 59 (19) |
| % over 65 | 51 (361) | 24 (875) |  | 19 (6) |
| % female | 59 (437) | 64 (2308) | <0.001 | 22/32 |
| % White British | 94 (714) | 88 (3142) | <0.001 | 28/32 |
| % with long term condition | 43 (298) | 51 (1747) | <0.001 | N/A |

For NHS 111 staff interviews, we interviewed commissioners (n=2), head of integrated and urgent care (n=1), clinical team manager (n=1), non-clinical team leaders (n=2), clinical advisers (n=6) and health advisers (n=4).

### Findings

The survey results have been published elsewhere (11) but have been summarised in table 1 below, to add context and explanation to the converged findings that are presented in this paper.

| **Services contacted in NHS 111 had not been available (excludes respondents who identified multiple services)** | **Telephone % (n=696)** | **Online % (n=2037)** | **P-value (unadj)** | **P value** |
| --- | --- | --- | --- | --- |
| Called 999 ambulance | 20 (138) | 1 (16) | <0.001 | <0.001, |
| Visited Emergency Department | 26 (181) | 3 (64) |  |  |
| Contacted Primary care (GP) | 29 (201) | 17 (341) |  |  |
| Contacted MIU/WIC/UCC | 18 (125) | 5 (100) |  |  |
| Phoned 111 | - | 21 (427) |  |  |
| Would not have contacted a service | 5 (35) | 43 (874) |  |  |
| Something else+ | 2 (16) | 11 (215) |  |  |
| **Helpfulness of the advice given by NHS 111** | **Telephone % (n=292)** | **Online % (n=3515)** | **P-value (unadj)** | **P value, chi-sq diff, adj\*** |
| Very helpful | 69 (202) | 43 (1495) | <0.001 | <0.001, 0.33 (0.24-0.44) |
| Quite helpful | 21 (61) | 32 (1138) |  |  |
| Not very helpful | 8 (22) | 15 (515) |  |  |
| Not helpful at all | 2 (7) | 10 (367) |  |  |
| **Satisfaction (% giving highest rating)** | **Telephone % (n)** | **Online % (n)** | **P-value (unadj)** | **P value, chi-sq diff, adj\*** |
| Overall, how satisfied or dissatisfied were you with the 111 service? ('very satisfied') | 71 (555) | 50 (1880) | <0.001 | <0.001, 0.40 (0.33 to 0.49) |
| How likely are you to recommend the NHS 111 service to friends and family ('very likely') | 69 (543) | 57 (2083) | <0.001 | <0.001, 0.55 (0.46 to 0.67) |
| How likely are you to use NHS 111 Online again ('very likely') | NA | 56 (2028) | NA | NA |
| The questions about the symptoms were relevant (strongly agree) | 51 (145) | 39 (1250) | <0.001 | <0.001, 0.55 (0.41 to 0.73) |
| **Compliance with the advice given by NHS 111** | **Telephone % (n=711)** | **Online % (n=3599)** | **P-value (unadj)** | **P value, chi-sq diff, adj\*** |
| Full compliance with advice | 88 (683) | 67.5 (2430) | <0.001 | <0.001, 0.07 (0.04-0.11) |
| Partial compliance with advice | 9 (63) | - |  |  |
| Did not comply with advice | 4 (28) | 32.5 (1169) |  |  |
| **Why advice given by NHS 111 was not followed (for those who did not follow advice)** | **Telephone % N/A** | **Online % (n=1169)** |  |  |
| Wanted to wait and see if symptoms changed |  | 27 (313) |  |  |
| Didn't want to waste anyone's time |  | 26 (307) |  |  |
| Unable to follow |  | 24 (276) |  |  |
| Symptoms improved |  | 18 (209) |  |  |
| Did not agree |  | 15 (174) |  |  |
| Couldn't get an appointment |  | 8 (96) |  |  |
| Did not understand the advice |  | 4 (52) |  |  |

+includes dentist, pharmacy, and other

We identified four key themes from our triangulated data analysis that helped to identify how people used the different 111 services, and the potential for channel shift.

#### 1. Telephone 111 was associated with higher user satisfaction than online interactions, which may be due to improved confidence and trust in the dispositions reached following health adviser interaction

Survey results showed that telephone users reported higher levels of satisfaction with the service than online users across all comparable measures (see Table 1). Telephone users were more likely to report that they were ‘very satisfied’ with the service and ‘very likely’ to recommend the NHS 111 service to family and friends, even after adjusting for age, gender, ethnicity and LTC (p<0.001). Telephone users were more likely to rate the advice given as ‘very helpful’ (69% (202/292) v 43% (1495/3515) p<0.001), while a quarter of online users rated the advice as not helpful, compared to 10% of telephone users. Telephone users were more likely to feel that the questions about symptoms were relevant (see table 1).

Within interviews, both staff and users highlighted problems with ambiguous and over-simplified question wording within OL111. This lack of flexibility in how questions were answered in the online system appeared to contribute to the lower confidence in the online advice than telephone advice, with the human interaction of the telephone call perceived as key to extracting the most appropriate and relevant information from the user. Almost half of the OL111 users interviewed felt that the questions were poorly worded, over-generalised or not relevant, which made them question the professionalism of the service and reduced their confidence in the advice given.

> *“I found it was very basic and for me that meant that I would not have been completely reassured with any conclusive advice it would have given because it seemed very generic, you know? And it’s like those horoscopes that come through the email. It just didn’t seem professional enough for me” [OL111 user 20]*
>
> *“Well, I always like to talk. Well, it’s always better cos you can, you get a rapport with someone and usually it’s relevant questions… normally you can tell almost immediately whether they’re trained well enough to do the job or not…I have confidence in people… I suppose it’s because I prefer dealing with people than machines” [OL111 user 22]*
>
> *Yes. They’ve put their, what they perceive to be their symptoms, their issues, into the online service, and then whatever the online service is then telling them to do, they’re phoning in to the service well it says I need to this, or it says I need to do that. So then they want, in my perception is, they then want the human contact. They want somebody to say yes that’s right, that’s what you need to be doing. Or well I’ll assess it and I’ll do this for you (Tel111 staff 07)*

Concerns about inappropriate and difficult to understand terminology were reflected by staff who reported spending more time with OL111 referrals due to the need to re-triage users who had misunderstood questions. The ability of health advisers to probe and obtain ‘softer’ information that would guide users down the most appropriate algorithm was seen as key to obtaining trustworthy outcomes by both users and staff.

> *When we speak to someone I can, as a nurse, I can question that. How bad is your chest pain? ‘Oh it’s really bad’. But I say ‘I’m talking to you and I cannot feel that you’re struggling for a pain that is very, very bad. Your voice is, you know, fluent, you’re not struggling, you’re not gasping for air. So pain that’s really, really bad will not make you sound so well. So are you sure that you have understood this question?’ (Tel111 staff O6)*
>
> *“it [online assessment] drove you down one straight route, you couldn’t necessarily pose questions, whereas with something like chat over the internet you can actually ask questions” [OL111 user 24]*
>
> *“it [the questions] was very black and white. There was nothing in the middle, it was very general-, it was almost like can you breathe or can’t you breathe. There was nothing in the middle. (…) specific enough, there weren’t enough options” [OL111 user 2]*

In particular, the telephone 111 service was perceived to be preferable for more complex or urgent problems when the human interaction could help direct the user to more appropriate advice, or intervene where necessary. The importance of human factors in understanding the complexity of healthcare problems was again highlighted as key, particularly for users with chronic conditions or multimorbidity, as the online questioning did not enable consideration of underlying problems that may impact on the problem being consulted about.

> *They [the online question] said ‘is this a new cough’ you know, or is it the old cough come back, you know just a bland question ‘is this a new cough?’ just doesn’t hack it, you didn’t know how to answer it. [OL111 user 25]*
>
> *It was a bit limited with regards to COPD. The questions didn’t go anywhere and I suspect probably there was not enough depth for it to have been terribly useful. I was feeling I knew more than the 111. [OL111 user 24]*
>
> *I had a sore throat, but I had no option to say that I’m asthmatic as well, which was the reason why I am worried [OL111 user 20]*

#### 2. NHS 111 online was felt to provide a useful and convenient adjunct to the telephone service

Despite the concerns highlighted above, OL111 was felt to add value to the healthcare system and most users (56%) reported they would be ‘very likely’ to use it again. Participants described OL111 as their preference for information or advice relating to what they perceived to be simpler, low acuity health problems such as stomach bugs and colds, or identifying the location of an emergency dentist, particularly when their primary care services were unavailable.

Although advice was less trusted, the speed and convenience also made it a preferred option for people with time pressures (e.g. those with young children). Both the lack of waiting time and the speed of the actual process of doing the assessment were key, particularly at a time when waiting times for telephone 111 precluded quick access to a telephone adviser, with some using both services simultaneously or doing an initial screening with OL111 before using the telephone.

> *let’s say I’m looking after my son or something else is going on in my life I get a phone call or something, I can stop [NHS 111 Online] at any time I want, I can restart it at any time I want so I like that kind of convenience.[OL111 user 31]*
>
> *I looked up online while calling 111 *laughs* and then I hung up, I said I’d do it online. I think it’s easier and faster. [OL111 user 13]*
>
> *…it’s it in some ways it’s better than the phone service, because you don’t have that waiting in a queue and if you’ve got a child and you’re worried about like I would be worried like my daughter with the swelling and her eye was closing up, I didn’t want to be sitting there for maybe half an hour or one in a queue looking an thinking, well is; should I be really be ringing for an ambulance you know is this gonna get worse. Erm so in that instance it was really good that I could just go on immediately, I was not in a queue, and I could just get advice, answer the questions and get advice, tailored to my answers really. (OL111 user 10)*

#### 3. NHS 111 online widens access to NHS 111 services for some subgroups of users who would not otherwise access the telephone service

Whilst the lack of personal contact was felt to negatively influence the quality of advice, it provided access to the 111 service for a subset of users who would not normally use the telephone service. This included people who did not like or were unable to use the telephone due to communication difficulties (e.g. hearing or language), or lacked a private space in which to talk. The impersonal nature of the interaction also suited people who wanted to avoid human interaction due to social anxiety or due to the sensitive nature of the problem being consulted about (e.g. genito-urinary, sexual or mental health problems).

> *I usually choose the online just cause it’s slightly easier because like I do have a bit of like anxiety issues, and like while I can talk on the phone and stuff, like a lot of the time it’s easier just to go online [OL111 user 18]*
>
> *If I don’t understand something then you know you never know how people explain things. Sometimes accents can be… like English accent…the pace someone is speaking [can be a problem]…factors like that are something that I would consider. So for me it’s only when I can have a think about or check the translation of some questions… it’s the time that I can take to do, to answer the questions. I feel, you know, on the phone, I would be stressed a bit just to answer as quickly as possible because there might be someone else waiting. So for me, the time I can do it and then having the option to go back to the questions [OL111 user 19]*
>
> *We get, we get erm, I suppose it’s surprising, for me alarming amount of people with mental health problems. Because that, in a way, it’s frightening because you don’t want them to just fill forms in, you want to be able to talk to them. But we do understand that, it might be a really good first step for seeing help. Because you don’t have to talk to somebody, you can just fill, fill those in, and then sort of cast it out and hope that someone can help you. Er, as opposed to ringing and having to, you know, say out loud how you feel. (Staff 01)*

Perceptions of OL111 as a ‘computer’ rather than human-based service enabled access for people who were conscious of not ‘over-burdening’ the healthcare system by contacting a health professional when they were unsure whether this was necessary. A quarter of survey respondents who explained why they did not comply with OL111 advice reported this being due to reluctance to ‘waste anyone’s time’ and concerns about burdening the health service were also reflected within user and staff interviews. Participants explained how they accessed the online service due to concerns about using health professional’s time inappropriately.

> *Well yes, because I’m going to be wasting their time, well let me rephrase that. I am going to be wasting other people’s time less […]. Unfortunately I’ve got mixed issues medically so I do need information sometimes where you know I don’t know what to do so for people like me, I think it’s actually a lifeline and saves all that hassle of wasting time going to the hospital or calling somebody in the middle of the night. [OL111 user 8]*
>
> *…I feel that you don’t seem to be wasting anybody’s time as much as if you were phoning up. (OL111 user 04)*
>
> *I think a first port of call to save taking up peoples time unnecessarily, erm that it’s a good place to go. If only to reinforce what you’re already thinking. [OL111 user 24]*
>
> *I also think it’s good for you know it puts less strain on the system obviously, than having to have staff man a phone, and so and that is, that is important like not just to the wider NHS but also to me personally Because you know when you feel like you’ve got something not that serious and you maybe don’t want to bother someone, erm that is a factor for me (OL111 user 31)*

#### 4. OL111 was perceived as more ‘disposable’ and used more speculatively by users but staff still felt responsibility for users contacting them for call back

The lack of human interaction and perceived lack of responsibility associated with OL111 interaction enabled people to use it more speculatively than the telephone service. Survey responses showed a significantly higher proportion of OL111 users saying they would not have contacted another service if they had not used 111, and also lower reported compliance with advice (68% reporting full compliance with advice for online users, v 88% for telephone users) with almost half of responses stating this was due to symptoms improving, or waiting to see if they improved.

Users described an implicit understanding of a hierarchy of services that they would use, with online 111 lower down the hierarchy than telephone 111. For many users, OL111 was seen as a limited version of Tel111 that offered ‘the next thing to Googling’ (OL111 user 20), providing some reassurance about decisions (due to the NHS trusted brand) about which service to use rather than definitive, trusted advice. Participants characterised the service as ‘just a computer’ and therefore limited in ability and usefulness.

> *This is more just generally concerned, but not too worried if I can put it that way, I might use the online service but if I had any genuine concerns, I would probably call 111 and if I had any real concern, I’d phone 999. (OL111 user 25)*
>
> *I: So, what, would you use the online again do you think?*
>
> *P: Erm I, I would but I wouldn’t rely on it, I’d probably look at it and then probably ring 111 telephone anyway. If I really needed proper advice. I, I, I would look at it out of interest if it’s something that’s not too bad and I might be thinking, ‘Oh what might this be?’ But I’m just as likely to look on the NHS website, cos that gives me stuff as well (OL111 user 03)*
>
> *It’s not so good so much for like possibly like I suppose, telling you what might be wrong with you. But then like I was saying, well it can’t really because it’s only a computer service (OL111 user 18)*

However, whilst users perceived OL111 as a low-resource and low-risk service, staff reported feeling a sense of responsibility and concern about patients who were referred into the telephone service by OL111. They characterised online interactions as ‘disposable’ and free from personal responsibility for patients, but felt a sense of responsibility and anxiety when they were unable to contact patients for a call-back, particularly when they had lower levels of trust in the data the user had entered as part of their online assessments.

> *So, if people fill them in late on, in the evening, we tend to be unable to make contact with them, erm, because they, they fill them in and then go to sleep [laughs]. And I think that although it tells them they’re gonna get a call back, I think like a lot of things that are online, there’s a sort of disposable element to it. They’ll fill it in at the time, because they’re interested in it, and then forget about it. (Staff 01)*
>
> *So if somebody says I’ve taken three more types of pain relief, and they’re not answering, then I’m worried because I’m thinking how much pain relief have they been taken? And then if they’re not answering again, I’m thinking am I safe to close this call or not? (Staff 06)*

## Discussion

### Summary of findings

Our findings suggest that human interactions involved in the telephone consultation appeared key to participants demonstrating higher levels of trust in telephone than online recommendations. Telephone users were more likely to rate the advice given as ‘very helpful’ and to be very satisfied with the NHS111 service, potentially due to telephone health adviser skills in probing and obtaining ‘soft information’ leading to more appropriate and trusted advice than online interactions. Users like the speed and convenience of NHS 111 online and perceived it as a useful and convenient adjunct to the telephone service. The lack of personal interaction led to widening access to NHS 111 services for some subgroups of users who would not otherwise access the telephone service. The nature of the online consultation meant that OL111 was perceived as more ‘disposable’ and used more speculatively.

Our findings suggested that perceptions of OL111 and Tel111 were shaped by perceptions of one as a human and the other as a computer-based service, with higher trust in the human. Staff and users described how the human interaction involved in the telephone call enabled them to provide more appropriate and accurate responses to the questions which in turn increased their confidence in the dispositions provided. Conversely, the perception of OL as a computer service, void of human interaction, opened up access to the 111 service for a subset of users who were drawn to the lack of human interaction, either because they found this difficult, or because they did not want to burden services involving a healthcare professional.

Previous literature reviews have highlighted the lack of evaluation of user experience of digital and online symptom checkers. (9, 13) Existing studies evaluating online and digital systems focus principally on areas of diagnostic accuracy and patient safety, (9, 14) with a lack of evidence comparing them to telephone triage services. (15, 16) (13) Compliance with advice was reported to be around 57% in a small web-based triage system in the Netherlands, (17) which was similar to our reported 67%. Lee et al (2022) reported user reluctance to trust health apps, compared to health professionals.(18)

There is some evidence that algorithm-based triage may be more risk averse than triage involving human interaction, with the potential to recommend higher levels of care than necessary. (19) Similarly, studies have reported diagnostic accuracy of online symptom checkers to be lower than triage systems involving humans. (16, 20). This supports the findings within our study that suggest human interactions are required to obtain more pertinent information than can be obtained via digital or online symptom checkers.

#### Strengths and limitations of this study

This study is the first to combine views of staff and users of the online 111 and telephone 111 services to understand how the services are being used, including results from a national online NHS 111 user survey. The use of a convergent parallel mixed methods design allowed us to converge findings from three different data sources to obtain a more in-depth understanding of the complexities of service use that may explain potential for channel shift.

The majority of the data collection undertaken for this project was undertaken prior to the Covid-19 pandemic. During the pandemic, in particular the first lockdown, demand for both online and telephone 111 soared, in part due to public health messaging asking people to contact 111 as a first point of call. During the pandemic there were long waits for the telephone 111 service, with recorded messages asking people to use the online service, which may have changed use of the online service since. Health-seeking behaviours may have changed as a consequence of the pandemic and it is likely that public perceptions of both online and telephone 111 will have changed since.

Use of online 111 and actions undertaken in response to the disposition provided were shaped by social expectations and protection of the NHS, which may not be transferable to health settings in other countries.

Although the response rate was low, it is similar to other surveys reported to the National NHS 111 minimum dataset. The characteristics of respondents was representative of the NHS111 telephone and online users in terms of gender of respondents, although younger people (under 35) were under-represented in our sample (11).

Due to the pragmatic need to use existing user surveys, user survey data compared two different populations (online and telephone), rather than asking the same group of people about both their experiences of both OL111 and Tel111. Differences in user satisfaction may therefore be partly due to differences in the populations surveyed. Adjekum et al (2018) identified that sociodemographic factors influence individual’s trust in digital health.(21)

## Implications

It is unclear whether there will be significant channel shift, particularly given how the two services are used differently. Online 111 seen as a useful adjunct to but not a replacement for telephone 111 and used for simple conditions, as a more trusted alternative to Google, but will potentially lead to induced demand rather than significant channel shift. This may have an impact on overall demand for wider services, but any increase in service use may be limited if users do not follow the advice provided. The telephone service provided higher levels of trust and satisfaction in both the interaction itself and the recommendation provided, which suggests that the telephone service would always be a preferred first option when call times allowed. If the online service did not provide users with what they needed, the induced demand may diminish over time.

Whether people use the online or the telephone service will be a function of the busyness of the telephone service and the reason for the call. For some, the online service will deliver all that they require, but we do not know what they would definitively have done without the online service. The telephone 111 service was perceived to be preferable for more complex or urgent problems. Future development of OLNHS111 will require refinement of algorithms, particularly for people with multimorbidity and long term conditions, perhaps alongside a live chat option to check that questions have been understood correctly. This could include asking whether they managed to say everything they felt was important with respect to their health problem.

Communication about NHS 111 telephone and online services should include advice about which service to use, and help to guide users towards the most appropriate service for their need at the first point of contact.

## Data Availability

All data produced in the present work are contained in the manuscript

## Acknowledgements

The authors would like to thank: all of the staff and 111 online and telephone users who gave their time to help with the research, Dan Fall and members of the Sheffield Emergency Care Forum (PPI group) for their useful feedback, NIHR for funding the research, members of the steering group and Marc Chattle for providing administrative support throughout.

## Ethical approval

Ethics approval for the interview-based work with service users and stakeholders was granted by North West Haydock Research Ethics Committee (reference 19.NW/0361). The University of Sheffield Research Ethics Committee granted ethics approval for the telephone and online NHS 111 user surveys (reference 030991)

## Author contributions

JT conceived the study, obtained research funding, oversaw the study and contributed to the analysis. EK co-ordinated the user surveys, undertook user interviews and led the analysis of user survey and interview data. JL undertook user interviews and contributed to the analysis of interviews. FS undertook staff interviews and analysis, led the converged analysis and drafted the article. All authors contributed to the research design. JC contributed to the analysis and drafting the paper. FS drafted the article and EK, JL, JT and JC critiqued the paper for important intellectual content. FS takes responsibility for the paper as a whole.

## References

1. NHS England. The NHS Five Year Forward View. NHS England: Leeds, 2014.

2. NHS England. Website 2022 https://www.england.nhs.uk/statistics/statistical-work-areas/ (accessed 24/11/2022).

3. Turner J CJ, Chambers D, Cantrell A, Phung VH, Knowles E. et al. What evidence is there on the effectiveness of different models of delivering urgent care? A rapid review. Health Serv Deliv Res. 2015;3(43).

4. NHS England. NHS 111 minimum data set 2018-19. 2018.

5. NHS England. Next steps on the NHS Five Year Forward View. NHS England: Leeds, 2017. 2017.

6. Carter M, Fletcher E, Sansom A, Warren FC, Campbell JL. Feasibility, acceptability and effectiveness of an online alternative to face-to-face consultation in general practice: a mixed-methods study of webGP in six Devon practices. BMJ open. 2018;8(2):e018688.

7. Cowie J, Calveley E, Bowers G, Bowers J. Evaluation of a Digital Consultation and Self-Care Advice Tool in Primary Care: A Multi-Methods Study. International journal of environmental research and public health. 2018;15(5).

8. Madan A. WebGP: the Virtual general practice. London; 2014.

9. Chambers D, Cantrell A, Johnson M, Preston L, Baxter SK, Booth A. Digital and online symptom checkers and assessment services for urgent care to inform a new digital platform: a systematic review. 2019;7(29).

10. Nijland N CK, Boer H, et al. Patient use and compliance with medical advice delivered by a web-based triage system in primary care. J Telemed Telecare. 2010; 16:8–11.

11. Turner J KE, Simpson R, Sampson FC, Dixon S, Long J, Bell-Gorrod H, Jacques R, Coster J, Yang H, Nicholl J, Bath P, Fall D, Stone T. Impact of NHS 111 Online on the NHS 111 telephone service and urgent care system: a mixed methods study. Health Serv Deliv Res. 2021;9(21).

12. Creswell JW, & Plano Clark, V. L. Designing and conducting mixed methods research 2011.

13. Gottliebsen K, Petersson G. Limited evidence of benefits of patient operated intelligent primary care triage tools: findings of a literature review. BMJ health & care informatics. 2020;27(1).

14. Wallace W CC, Chidambaram S, Hanna L, Iqbal FM, Acharya A, Normahani P, Ashrafian H, Markar SR, Sounderajah V, Darzi A.. The diagnostic and triage accuracy of digital and online symptom checker tools: a systematic review. NPJ Digit Med 2022;17(5(1):118).

15. Donovan E, Little P, Willcox ML, Wilcox CR, Patel S, Hay AD. Digital interventions for parents of acutely ill children and their treatment-seeking behaviour: A systematic review. British Journal of General Practice. 2020;70(692):E172–E8.

16. Yu SWY, Ma A, Tsang VHM, Chung LSW, Leung SC, Leung LP. Triage accuracy of online symptom checkers for Accident and Emergency Department patients. Hong Kong Journal of Emergency Medicine. 2019.

17. Semigran HL LD, Nundy S, et al Comparison of Physician and Computer Diagnostic Accuracy. JAMA Intern Med. 2016;176(1860-1).

18. Lee VV VS, Lau NY, Blasiak A, Siah KTH, Ho D.. Understanding the user: Patients’ perception, needs, and concerns of health apps for chronic constipation. DIGITAL HEALTH. 2022(8).

19. Hill MG, Sim M, Mills B. The quality of diagnosis and triage advice provided by free online symptom checkers and apps in Australia. The Medical journal of Australia. 2020.

20. Berry AC, Berry NA, Wang B, Mulekar MS, Melvin A, Battiola RJ. Symptom checkers versus doctors: a prospective, head-to-head comparison for cough. 2020;14:413–5.

21. Adjekum A BA, Vayena E.. Elements of Trust in Digital Health Systems: Scoping Review. J Med Internet Res. 2018; Dec 13;20(12):e11254.

